# Implementation of Robson classification for caesarean section using health insurance claims: the experience of Indonesia

**DOI:** 10.1101/2024.04.18.24306050

**Authors:** Levina Chandra Khoe, Euis Ratna Sari, Dwirani Amelia, Tauhid Islamy, Amila Megraini, Mardiati Nadjib, Budi Wiweko, Indah Suci Widyahening

## Abstract

Robson classification has been recommended by the World Health Organization (WHO) as a monitoring tool for caesarean section (CS), however, it has not been implemented in Indonesia. In this study, we hypothesize that the National Health Insurance (NHI) claims data can be used to classify pregnant women into several obstetric groups. This study aims to examine the use of NHI claims database for analyzing CS according to the WHO manual for Robson classification. This study is a cross-sectional analysis using delivery claims from NHI sample set data from 2017 to 2018. We categorized the International Classification of Diseases (ICD) 10 codes in the claims data according to the Robson classification system using the following variables: multiple pregnancy, fetal presentation, previous obstetric record, previous CS record, gestational age, and onset of labor. Data was analyzed using IBM SPSS Statistics. A total of 31,375 deliveries were included in the analysis. Overall, mean age of mothers was 29.2±5.9 years. The CS rate in this population was 37.0% in 2017 and 38.7% in 2018. Highest CS rate was found in nulliparous (group 2: 26.6%) and multiparous women (group 4: 24.8%) if labour induced or had prelabour CS, followed by multiparous women with previous uterine scar (group 5: 22.5%). We found an alarmingly high rate of CS among Indonesian women. Implementation of Robson classification in the National Health Insurance claims data is feasible and should be considered by the policy makers as an audit tool to identify the groups that contributes the most to the CS rate.

## Introduction

Historically, caesarean section (CS), a procedure to deliver baby through surgery, was performed as an attempt to save mothers and newborns [1]. However, in modern days, deciding on delivery mode is no longer determined merely by medical indications, but include non-medical reasons, such as mothers worried about normal deliveries, families who cannot bear to see mothers in pain, or limited support for vaginal birth [2–4]. With the increase in the variety of reasons for CS, it is not surprising that the number of CS has raise steadily. Globally, the rate of CS has increased 19.4% from 1990 to 2018 [5]. Similar trend happens in Indonesia, where the CS rates have risen from 10% in 2007 to 17% in 2017 based on the population survey [3]. Report from the medical claims showed an even higher rate of CS, i.e., 34.8% after the implementation of National Health Insurance in 2014 [6]. Both figures indicate higher rate of CS above the optimum range of 10-15% according to the World Health Organization (WHO) [6].

In 2001, Robson introduced the classification system of CS rates based on the obstetric characteristics of women, such as the category of pregnancy, previous obstetric record, the course of labour and delivery, and the gestation of pregnancy[7]. This classification system allows comparison of CS rates across different institutions and countries, as recommended by the WHO[8]. It has also been reviewed as the most appropriate classification system to identify why and on whom CS are being performed [9]. Few studies had been performed to assess the CS rate in large hospitals in Jakarta and Bali. They found that the CS rate in one hospital in Jakarta was 48.0% and in Bali 34.3%, much higher than the national average, i.e., 17.7% [10–12]. The most common indication for CS was preterm pregnancies, followed by malpresentation, and CS in previous pregnancy [10, 11].

Though, in 2018, the Basic Health Research (Riskesdas) has presented the CS rate at population level (17.7%), there is a paucity of data on indications for caesarean in Indonesia. It is difficult to draw conclusions regarding why CS rate is increasing and what strategies should be implemented to control this phenomenon. Alternatively, routinely collected data, such as insurance claims data, contains information on the pattern of obstetric service delivery. Data recorded in the National Health Insurance (NHI) claims only includes those who are members of the insurance, and it already covers more than 90% of total Indonesian population. Exploration of these rich data will be useful to identify which subpopulation is driving the high CS based on the Robson classification systems. Our study aims to assess the CS rate at national level, to identify the contribution of specific obstetric population to CS rate according to Robson classification, and to examine the use National Health Insurance claims database for analyzing CS rate according to Robson classification. The information derived from this study would be valuable to inform policy makers for strategies to monitor the CS rate across different hospitals in Indonesia.

## Methods

### Study design

This is a cross-sectional study, carried out through review of the National Health Insurance (NHI) claims sample set from 2017 to 2018.

### Data source

In 2020, the Social Security Agency for Health (BPJS-K) published a sample set of NHI claims data that could be accessed by the public. The source of this data comes from health facilities, both at primary and secondary level. The BPJS-K has selected approximately one percent of all members in accordance with proportionate stratified random sampling from the database. The samples selected from the database were individuals who are newly registered as NHI members in 2017 and 2018 with total members of 16,147,772 (2017) and 16,616,415 (2018). Family is used as a sampling unit. The strata used in the sampling method is developed based on combination of two variables: (1) unit of primary health care and (2) family category. Data was classified into three family categories: (1) families who never utilized any healthcare services; (2) families who utilized the primary care; and (3) families who utilized both primary and secondary care. This selection process resulted in a sample of families 56,791 units (2017) and 60,164 units (2018). The next step of sampling method is to select the individuals in each sampling unit. This step produces a total 134,966 individuals (2017) and 139,326 individuals (2018). For this analysis, we included all women in the International Classification of Disease, 10^th^ Revision, Clinical Modification (ICD-10 CM) in the code range O00-O9A for pregnancy, childbirth, and the puerperium.

### Robson classification system

We used the Robson classification systems as recommended by the WHO and the International Federation of Gynecology and Obstetrics (FIGO). The women in our sample set are categorized into one of ten groups classification according to their obstetric characteristics. The detail in each group is described in Table 1.

**Table 1.** Ten Groups Classification System.

| Group | Description |
| --- | --- |
| **1** | Nulliparous women with a single cephalic pregnancy, $\geq 37$ weeks gestation in spontaneous labour |
| **2** | Nulliparous women with a single cephalic pregnancy, $\geq 37$ weeks gestation who either had labour induced or were delivered by caesarean section before labour |
| 3 | Multiparous women without a previous uterine scar, with a single cephalic pregnancy, $\geq 37$ weeks gestation in spontaneous labour |
| 4 | Multiparous women without a previous uterine scar, with a single cephalic pregnancy, $\geq 37$ weeks gestation who either had labour induced or were delivered by caesarean section before labour |
| 5 | All multiparous women with at least one previous uterine scar, with a single cephalic pregnancy, $\geq 37$ weeks gestation |
| 6 | All nulliparous women with a single breech pregnancy |
| 7 | All multiparous women with a single breech pregnancy, including women with previous uterine scars |
| 8 | All women with multiple pregnancies, including women with previous uterine scars |
| 9 | All women with a single pregnancy with a transverse or oblique lie, including women with previous uterine scars |
| 10 | All women with a single cephalic pregnancy $< 37$ weeks gestation, including women with previous uterine scars |

### Variables

The dataset contains the demographic characteristics (e.g., age, gender, marital status, type of membership), primary and secondary diagnoses identified by the International Classification of Diseases (ICD) codes. All personal information has been removed to protect the individual’s privacy. Currently the NHI has not implemented the Robson ten-groups classification. Therefore, we develop a novel procedure to make the classification based on the ICD-10 codes as shown in Fig 1. The variables necessary for applying the Robson classification in the claims data are as follows: 1) multiple pregnancy; 2) fetal presentation (transverse/oblique/breech/cephalic); 3) previous obstetric record (nulliparous/multiparous); 4) previous CS record (with / without uterine scar); 5) gestational age (< 37 weeks / ≥ 37 weeks); and 6) onset of labor and delivery (induced/CS before labor/spontaneous).

**Fig 1.**
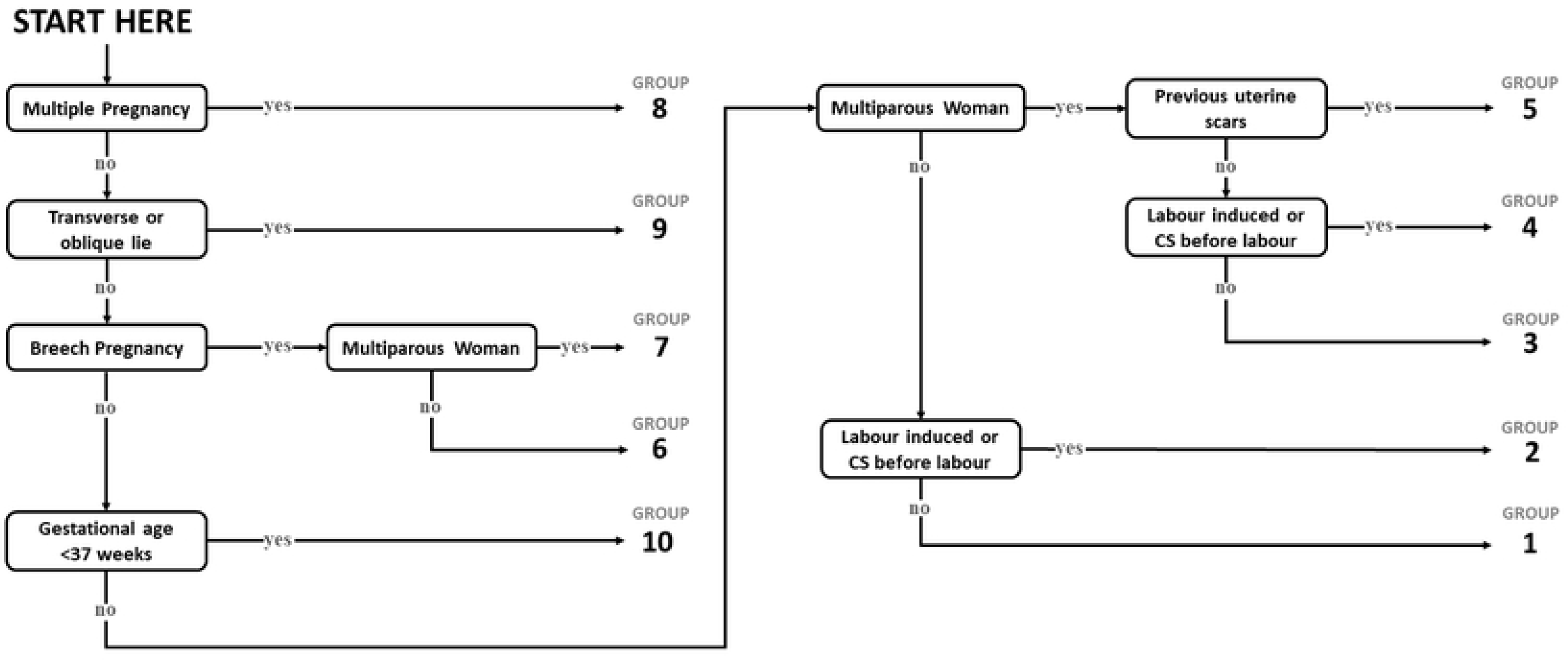
Classification of Women in the Robson Classification using ICD-10 codes. Source: Modified from the flowchart for the classification of women in the Robson Classification[8]

### Ethics

This study was approved by the Health Research Ethics Committee, University of Indonesia and Cipto Mangunkusumo Hospital number KET-472/UN2.F1/ETIK/PPM.00.02/2020. Data used in this study was obtained from the sample claims data, provided by the Indonesian Social Health Security Agency (BPJS Kesehatan) for the purpose of this study in June 2020. The data contains no personally identifiable information (de-identified data).

### Data analysis

Data was exported to and analyzed using IBM SPSS Statistics version 24 (IBM Corp., N.Y., USA). We reported the characteristics of women and calculated the overall CS rate by dividing the total number of cesarean deliveries with the total number of births. The women were categorized into one of the ten Robson groups according to the coding in Figure 1. For each group, we estimated the CS rate and its contribution to the overall CS rate. We used the WHO Robson implementation manual to interpret the results.

Before interpreting the CS rate, we assessed the quality of data and the type of obstetric population. The WHO suggests that the size of group 9 should be less than 1%[8]. If it is higher, it is probable that there were some misclassifications in this group. The CS group in group 9 should be 100%, if it is not, then it is assumed that there could be also a misclassification. We assess the type of obstetric population by comparing the size of each Robson group with the Robson guideline[8].

## Result

### Characteristics of women

A total of 31,375 deliveries were analyzed in this study. The mean age of women were 29.2 years (SD 5.9). The mean age of women with CS deliveries were significantly higher than women with vaginal birth deliveries, i.e., 29.7 years (SD 5.8) versus 28.5 years (SD 6.0) (p < 0.05). The characteristics of women are given in Table 2.

**Table 2.**
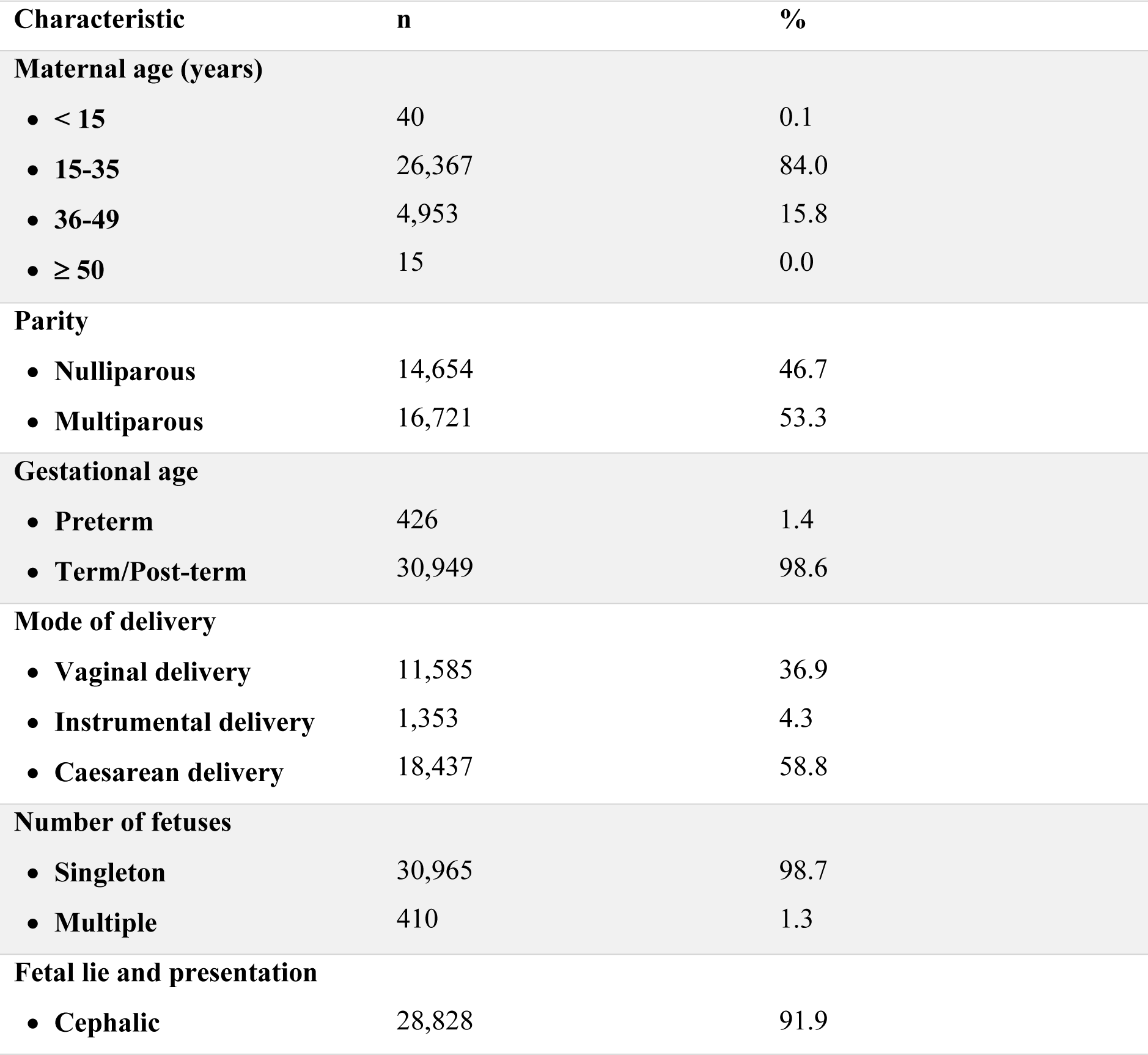
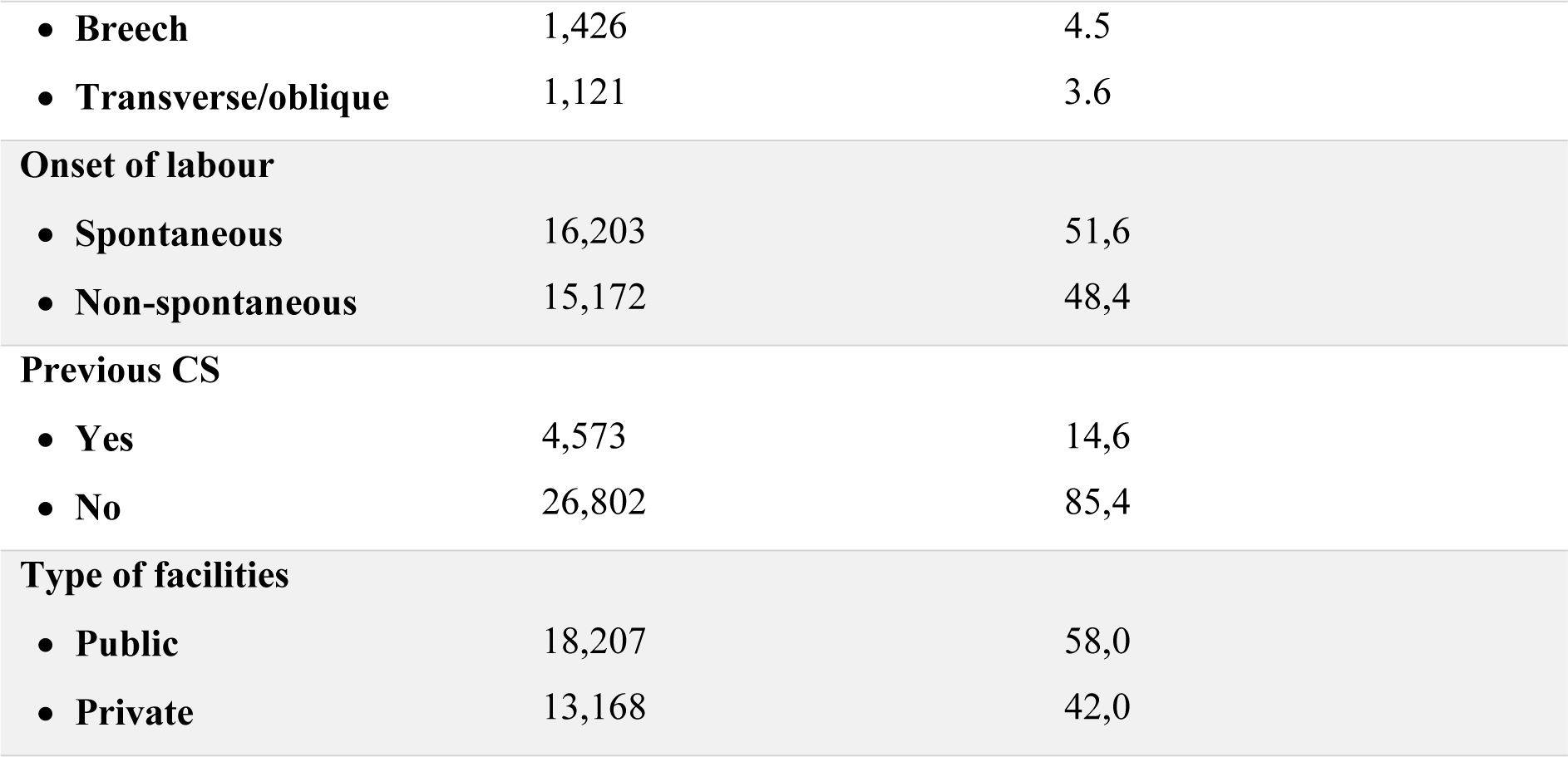
Characteristics of women in the Indonesian National Health Insurance (NHI) claims sample set (2017 to 2018; n total = 31,375).

### CS rate in each group

In terms of the group size, women in the group 3 made the largest contribution to the overall obstetric population, i.e., 22.2%. This was followed by group 1 and group 2, which accounted for 18.9% and 17.7%, respectively. The proportion of CS among all deliveries was 37.0% in 2017 and 38.7% in 2018, consecutively. Using the Robson classification system, we identified the largest contributing group to the overall CS rate was group 2 (26.6%), followed by group 4 (24.8%), and group 5 (22.5%). The distribution of CS delivery is provided in Table 3.

**Table 3.** Distribution of CS delivery in the Indonesian National Health Insurance (NHI) claims sample set (2017 to 2018) according to the Robson classification system (n total = 31,375).

| Group | Group size (%) | Group CS rate (%) | Absolute group contribution to overall CS rate (%) | Relative contribution of group to overall CS rate (%) |
| --- | --- | --- | --- | --- |
| 1 | 18.9 | 18.9 | 3.6 | 6.1 |
| 2 | 17.7 | 87.9 | 15.6 | 26.6 |
| 3 | 22.2 | 15.8 | 3.5 | 6.0 |
| 4 | 17.1 | 85.4 | 14.6 | 24.8 |
| 5 | 13.6 | 97.2 | 13.2 | 22.5 |
| 6 | 2.0 | 78.1 | 1.6 | 2.7 |
| 7 | 2.5 | 76.9 | 1.9 | 3.2 |
| 8 | 1.3 | 76.6 | 1.0 | 1.7 |
| 9 | 3.5 | 98.5 | 3.5 | 5.9 |
| 10 | 1.2 | 28.9 | 0.4 | 0.6 |

Note: Group size (%) = n of women in the group / total N of women delivered in the hospital x 100; group CS rate (%) = n of CS in the group / total N of women delivered in the group x 100; absolute group contribution to overall CS rate (%) = n of CS in the group / total N women delivered in the hospital x 100; relative contribution of group to overall CS rate (%) = n of CS in the group / total N of CS in the hospital x 100

## Discussion

More than one-third of women who delivered at hospitals in Indonesia went through CS procedures (37.0% in 2017 and 38.7% in 2018). These high CS rates have raised a question whether this CS procedure was deemed necessary and appropriate according to medical indications. This number is much higher than the average CS rate at global level (19.9%)[13] and among other South-east Asian countries (1.51%-31.78%)[14]. Nevertheless, the result should be interpreted cautiously since it did not reflect CS rate at population level, and merely included deliveries at secondary care level and women who were members of NHI.

Before we could interpret the CS rate in each group, we should assess the quality of data and type of obstetric population. The WHO suggests that the size of group 9 should be less than 1%[8]. However, we found in this study that it was more than 1%. It is probable that women with breech were misclassified to be allocated into transverse/oblique group. The CS group in group 9 should be 100%, but we found the CS rate was 98.5%. Thus, we assumed that there could be a misclassification in the categorization of Robson group.

The next step is to assess the type of obstetric population. The size of group 1 and 2 (nulliparous women) in this study was 36.6%. It was still in accordance with the Robson guideline, i.e., 35-42%[8]. While the size of group 3 and 4 (multiparous women) was 39.3%, higher than the recommended percentage (30%)[8]. The size of group 5 (previous CS) was 13.6% - related to the CS rate among nulliparous women in the previous year. Considering the overall CS rate was high, it is acceptable that the size of group 5 will also be high (more than 10%). The size of group 6 and 7 was 4.5%, slightly higher than the recommended percentage (3-4%). The size of group 8 (multiple pregnancies) was 1.3%, which is still appropriate. The size of group 10 (preterm) was 1.2%, it is within the guidelines (<5%). The ratio of the size of group 1 versus group 2 is around 1.

Multiparous women without previous caesarean section (Robson group 3) made up the biggest portion of all obstetric population in Indonesia. With over than 270 million population and rank 4^th^ as the largest population in the world, the Indonesian’s fertility rate is 2.3 live births per women, higher than its neighboring countries, like Singapore, Malaysia, and Thailand. Yet, in comparison with other countries with similarly low Human Development Index (HDI), the size of multiparous women in Indonesia is lower, 22.2% versus 43.7%[15]. These might have been associated with the implementation of family planning in Indonesia, with contraception prevalence rate has increased from 40% (1990) to 63% (2017) and the unmet need for family planning has reduced from 17% (1991) to 10.6% (2017). The contribution of this group to overall CS rate is considerably low, which means more women in this group delivered through spontaneous labor.

The highest contributor to the overall CS rate were nulliparous women (group 2) and multiparous women (group 4) who either had labor induced or were delivered by CS procedure before labour. Almost all women (> 85%) belong to these groups would end up with CS procedure. The result was higher than a WHO study in 21 countries and a study in two hospitals in Brazil that found less than 10% of CS rate in group 2 and 4[15, 16]. Antenatal care is essential for pregnant women to provide screening and tests to detect high risk pregnancy[17]. In Indonesia, every pregnant woman is recommended to at least have four antenatal care visits during their pregnancy: one visit in the first trimester, one visit in the second semester, and two visits in the third trimester[18]. Four antenatal care visits as suggested by the Indonesian Ministry of Health is in line with the WHO antenatal care model in 2002[19]. The standard of Indonesian antenatal care covers the following procedures, such as measuring mothers’ weight, upper arm circumference, fundal height, monitoring blood pressure and fetal heart rate, identifying fetal presentation, administering tetanus toxoid immunization, providing iron tablets, conducting laboratory tests, providing health education, and making appropriate referrals[20]. The new guideline by the WHO suggests more frequent antenatal care visits, i.e., 8 visits, for a positive pregnancy experience[21]. Unfortunately, even using the previous WHO manual, there were about 25% of pregnant women who had not completed the four antenatal care visits [22]. Additionally, many still did not have good maternal knowledge, including the knowledge on pregnancy emergency sign, sign of childbirth, preparation for complications and childbirth[23].

Women who experienced CS procedure in their previous pregnancy (group 5) also made substantial contribution in the overall CS rate. Our findings showed that the CS rate in this group was higher than the WHO study, a study in Canada and one tertiary hospital survey in Indonesia [15, 24, 25]. The high CS rates in group 1 (nulliparous women in spontaneous labor) could drive the increase rate of CS in subsequent pregnancies (group 5) [26]. Factors that might be associated with the increase CS rate in the first pregnancy are advanced maternal age[27], obesity[28], hypertension during pregnancy[29], fear of pain in labor, convenience[30], physician-induced demand[31], and inappropriate maternity care[32, 33]. In our study, only 15.8% of the mothers were in the age group above 35 years old. Other factors could not be identified in the claims data.

By having CS in the first pregnancy, women have higher chance to experience another CS[34]. Vaginal birth can be a safe option for women with previous cesarean delivery[35]. The success rate of vaginal birth after caesarean section (VBAC) varies from one study to another, ranging from 50%[36] to 85%[35]. Despite the high success, the utilization rate of VBAC is still low. In United States, only 8.2% women went for VBAC in 2007[37]. However, there is no data currently available to reflect the utilization rate of VBAC in Indonesia. The low VBAC rate has been assumed to associate with concerns on VBAC complications, such as uterine rupture, blood transfusion, puerperal sepsis, and surgical injury[38]. Additionally, the low rate is influenced by the women’s lack of knowledge on VBAC, fear of childbirth, and physician’s fear on the medico-legal liability[39, 40]. There are also concerns on the lack of skills and infrastructures, particularly in area with low resources [32, 41, 42].

To our knowledge, this is the first study that investigate the caesarean deliveries using Robson classification at the national level. Since Indonesia does not have an integrated medical record systems across hospitals, the use of claims data to reflect the CS rate at national level and monitor the CS trend at hospital and province level should be considered in the national policy, as the implementation is feasible as shown in our study. However, we are aware of the limitation in this study, such as the possible of misclassification of women into one of the Robson groups. The CS rate in group 9 was higher than WHO recommendation, and this could indicate poor data quality. Further modification in the coding system should be done to monitor the obstetric practices in all hospitals in Indonesia. Additionally, our study only reflected the CS rate at secondary care level, but unable to provide findings at primary care level.

## Conclusion

Using claims data, we conclude that more than half of women who delivered in the hospitals went through CS procedure. This study provides a preliminary investigation to assess the use of Robson classification system to identify which obstetric groups that contribute to the CS rate at national level. Our findings showed that implementing Robson classification in the claims data is feasible and useful to present evidence at hospital, regional, and national level. Therefore, we suggest embedding the Robson classification in the National Health Insurance claims application, thus, it will be able to identify the potential of unnecessary CS procedure.

## Data Availability

The sample set of National Health Insurance (JKN) data is available for public upon reasonable request to the Indonesian Social Health Security Agency (BPJS Kesehatan).

## Acknowledgements

Not applicable

